# Playing the Harmonica with Chronic Obstructive Pulmonary Disease. A qualitative study

**DOI:** 10.1101/2021.08.01.21261448

**Authors:** A Lewis, J Conway, J Middleton, C Startup, J Wyatt

## Abstract

**Introduction:** Pulmonary Rehabilitation (PR) is the gold standard, group-based intervention for individuals with Chronic Obstructive Pulmonary Disease (COPD). However, accessibility and adherence to PR is sub-optimal. Arts in Health interventions also improve health outcomes for people living with long term conditions. Playing the harmonica with COPD could be clinically beneficial. However, little is known about the patient experiences of playing the harmonica.

**Methods:** A qualitative, interpretivist, phenomenological study was undertaken, exploring COPD patient experiences of harmonica playing with a group of others living with chronic respiratory disease. Semi-structured interviews were completed, transcribed, and reflexive inductive thematic analysis performed.

**Results:** Eight people with COPD were interviewed. Thematic analysis generated five themes. Themes included “Hard in the beginning”, “Holding the condition”, “Breathing control”, “Gives you a high” and “Needing the Zoom class”. Playing the harmonica with COPD is difficult at first, particularly drawing a breath through the harmonica. With practice, experience in a fun activity, and quality teaching, individuals were able to become more attuned and embodied with their breathing. As breathing became easier the songs, rather than breathing, became the focus, and participants were able to escape living with respiratory disease when playing. The group was a priority in the weekly lives of participants, even though the buzz of being part of a group was lost.

**Discussion:** Playing the harmonica requires a different way of breathing and offers a breathing control strategy. Participants also reported the harmonica helped airway clearance and enabled a continued, regular social interaction through COVID-19. The results of this study compliment previous quantitative results and are relevant to physiotherapy. Further mechanistic studies and randomised controlled trials are needed to investigate the biopsychosocial benefits of playing the harmonica with COPD.

## Introduction

Chronic Obstructive Pulmonary Disease (COPD) is one of the largest causes of morbidity and mortality globally(1). Evidence-based interventions are available to help treat and manage disease burden, such as flu and pneumonia vaccinations, smoking cessation, inhaled therapies, and lung volume reduction(2). Pulmonary rehabilitation (PR) is a valuable intervention to improve symptom burden, exercise capacity, and quality of life (3, 4). Pulmonary rehabilitation also enables individuals with similar conditions to socialise and learn from one another, which is important considering the detrimental effects of social isolation to functional health status when living with COPD(5). However, only the minority of eligible individuals with COPD are referred and complete PR(6).

Arts-in-health interventions are recognised as being beneficial for health outcomes by the World Health Organisation(7). Singing for Lung Health (SLH) is an arts-in-health intervention with evidence for improving physical health in respiratory disease(8). Other arts-in-health interventions such as dancing, and other music therapies maybe appropriate intervention choices for individuals with chronic lung diseases(9-11). The harmonica is a hand-held instrument requiring the player to draw and blow against resistance in order to produce a tune, and has potential as an arts intervention for people living with COPD. Hart et al(12) performed a cohort study with individuals with COPD who participated in 12 weeks of harmonica sessions. These individuals improved their respiratory muscle strength (PImax mean difference = 15.4 cm H20, PEmax mean difference = 14.4 cm H20) and walking distance (6 minute walk test mean difference = 61 metres), but there was no control group for comparison. In a small controlled study, Raharjo et al (13) explored the effectiveness of weekly harmonica sessions for 6 weeks for 34 individuals with COPD. At baseline there were no significant differences between treatment and control groups in inspiratory capacity, functional breathlessness, walking distance or quality of life. Harmonica playing improved inspiratory capacity (0.54±0.30 litre) and 6 Minute walking distance (90 ±23.96 meters), decreased mMRC (breathlessness) score (2.00±0.48) and St Georges Respiratory Questionnaire score (Disease specific quality of life) (32.35±5.57). All these changes are clinically relevant. However, when harmonica playing has been trialled in combination with PR compared to participating in PR only, no statistically significant differences in respiratory muscle pressures, exercise capacity or quality of life were observed(14). It is not known what the experience of playing the harmonica is like for individuals living with COPD. The objective of this research was to investigate the experience of playing the harmonica for individuals who are part of a harmonica group specifically created for people living with chronic respiratory disease. Experiences of being part of a group included both face-to-face and remote group delivery.

## Methods

This was a qualitative study using an interpretivist, phenomenological stance. AL is a respiratory physiotherapist, has a background in pulmonary rehabilitation, and has previously experienced SLH groups. He has tried playing the harmonica on multiple occasions, but does not play as a regular hobby. The phenomenology was influenced by Merleau-Ponty regarding corporeality, with the body as a perceiver and actor (15) and also by Van Manen (16) with a view to practice, with the intent of caring for individuals with respiratory disease.

Individuals were recruited from a harmonica group in the UK, consisting of individuals living with different chronic respiratory diseases. Participant information sheets were given to all individuals in the group diagnosed with COPD who had participated in at least six weekly sessions. Interested individuals then contacted AL for participation in the research and provided informed consent. AL had no previous relationship with recruited participants. Ethical approval for the project was obtained by Brunel University London College of Health, Medicine and Life Sciences Research Ethics Committee (25578-MHR-Dec/2020-29450-2).

Semi-structured interviews were performed via Zoom or telephone depending on participant preference. The semi-structured interview guide is provided in the online supplement. Interviews were transcribed verbatim and reflexive inductive thematic analysis was performed(17, 18). Respondent validation or peer checking of themes were not performed because this was deemed not appropriate for the interpretivist nature of the research. The harmonica group leader assisted in recruitment but was not involved in the analysis of the interviews.

The harmonica group was set up in 2017 as a follow-up session of an established SLH group. Sessions were run by an occupational therapist who is also a musician, and has previously been trained to run groups specifically for individuals with chronic respiratory disease (CRD) by the British Lung Foundation. All weekly harmonica sessions consisted of warm-up exercises and performing different songs specifically chosen by the singing leader to be appropriate for individuals who may become breathless. Harmonica sessions lasted one hour per week. All harmonica sessions were run via Zoom at the time of recruitment because of COVID-19 restrictions.

### Patient and public involvement statement

Patients and public were not involved in the design of this study. However, the patient experiences reported within this study will be considered by the research team in planning future studies. The patients involved in this study will be provided with a presentation of the results in a group setting following publication.

## Results

Out of a regular harmonica group attendance of thirty individuals, 15 had known COPD and were approached to enter the study. Eight participants (six females, two males) were recruited. Interviews lasted an average of 49 minutes. Five themes were generated from thematic analysis which included “Hard in the beginning”, “Holding the condition”, “Breathing control”, “Gives you a high” and “Needing the Zoom class”. The themes in bold text and subthemes in red text are presented in the thematic map figure 1 below. Further example quotes are provided in the online supplement.

**Figure 1:**
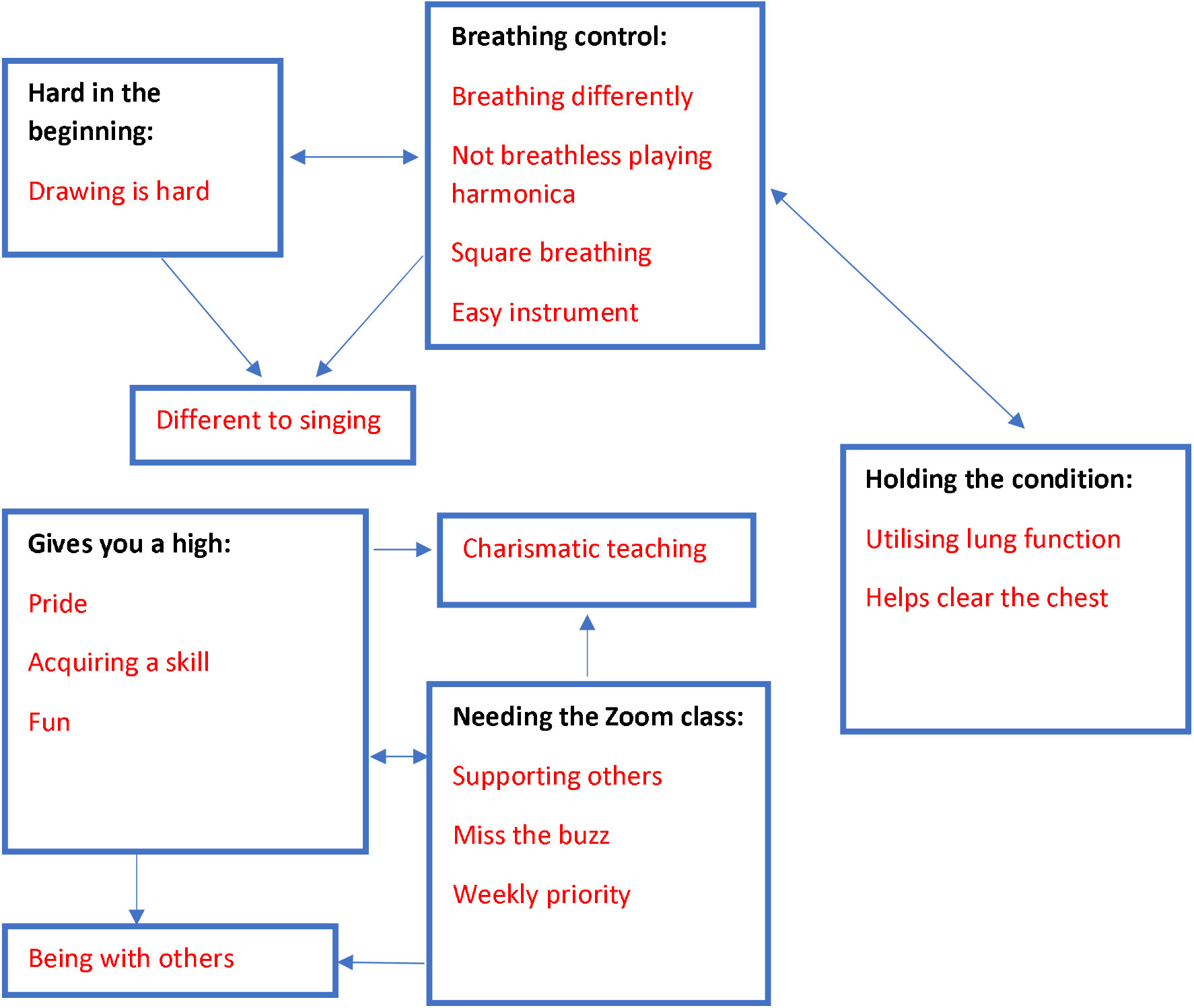
Thematic map

### Hard in the beginning

Participants reported that altering their breathing in combination with trying to hit the right note, was difficult. Many found that they were putting too much effort in to make the sound.

“first you find it hard but then once you get to realise how to breathe properly you’re actually playing a song without realising and you are not feeling breathless afterwards”

P1, page 3, line 53

“When I first started and found it so hard I couldn’t breathe to get enough breath to blow the harmonica, until I was shown how to do it.”

P2, page 3, line 8

The main difficulty was drawing the breath on the inhale to overcome the resistance of the harmonica:

“Blowing is okay, it’s drawing, and if you’ve got to do it very quickly, you can’t really take a good breath you know what I mean, when you’re breathing, in an out, it is much harder to do it through the harmonica. I don’t know whether that’s psychological or not because you’re still getting that breath in but I do find it can be quite strenuous and quite, if you’ve got to work at it to get in, to get the different drawing in and the blowing out you know, I have to concentrate and I have to think what I’m doing with the old lungs, you know it’s good for me to exercise them like that.”

P6 page 3 line 58

“because I’m drawing too much in at once, or I’m not breathing out enough because you’ve got to breath out to draw in.”

P8 page 4 line 86

### Holding the condition

Participants reported that they felt playing the harmonica regularly was a way that they were able to give their lungs a work-out and hold the condition, preventing them from worsening.

“I think it’s the structure that he puts in it, and he tells us you know how to do it, and to breathe, and to get the best out of our breathing. But I honestly don’t think I would be in the position I am now if I was still coughing and only on the medication that I was on, I think I would be in pretty dire straits”

P6 page 8 line 174

“It’s just making me breathe in a different way to what I would if I wasn’t doing it. It’s making me exercise when perhaps I wouldn’t do those exercises, because you know when I take my ventolin in the morning, because of harmonica, I now do their exercises for breathing and that helps me take my ventolin in my inhaler in the morning, so I just take one in the morning that helps me. Because you’ve gotta build up your lungs and capacity because you’ve lost you know, you’re losing it so I want to keep it and I can’t get what I’ve lost, that, but I want to keep it at the level now I am at the moment. I don’t wanna have to go onto oxygen like some of the ladies have, so I’m fighting it.”

P8 page 9 line 211

Participants reported that playing the harmonica helped to clear mucous off of their chest which helped in holding the condition on a day-to-day basis:

“I find it easier as of, sort of, forget I’ve got any chest problems when I’m playing the harmonica. I find the trainers and the flutter not as easy for me, I’m on the very, the thing with the spring, I’m on the very low level, I can’t seem to get up, so it’s strange that, put the harmonica into my lips and I’m fine, yes I do get out of breath (if) I played a lot, but that’s to myself. Because I play different things to the class but it’s not, I feel like it’s not a chore, the flutter and this the other one, I find I don’t do them regularly. I probably don’t need to because of using that harmonica.”

P4 page 8 line 184

### Breathing Control

A sense of needing to control the breath to play the harmonica or gaining a sense of controlled breathing through playing the harmonica, was a central theme in this study. Playing the harmonica was thought of as breathing differently, and often a way to forget about being breathless at all.

“it’s a lot lighter, it’s like I said before, it’s slightly whispering. It’s like you whispering in, it’s like you’re actually whispering into your harmonica so it’s not as hard….when you realise it’s coming, just coming from your throat area, and not down here, you tend to just blow into it gently.”

P1 page 5 line 110

“I forget that I have breathing problems when I do it, when I play, that’s what it means to me.” P3 page 14 line 321

“it is just asking you to be able to control your breathing, breathe at the right time and, and, and just produce the notes that you know the tunes that he wants you to play. And make sure that you’re playing the right notes of course. And it makes it quite simple for us because the notes not written as music notes, the written, his numbers, and are either, got a signal for blowing, or a signal for drawing, you know, so, so it makes it quite simple to do.”

P2 page 4 line 91

These quotes possibly indicate that through music, individuals are becoming more embodied with their breathing in a task where the product of action is not their breath, but the tune. This tune has no disease attributed to it and so participants lose focus on their own disease to some extent.

“It’s making your head work use your lungs, and I think that’s what we tend to miss out, that connection, when you panic, and you can’t breathe, and you panic and that’s it. So there’s no control up here at all, it is just one overriding panic, so doing anything that can help you ease that control and use that control, it’s got to be a fantastic thing for anybody with their respiratory problem.”

P5 page 19 line 441

“You’ve got to get your breath right in it, when you’re following the notes. Because it’s so easy to go off to get the wrong note. So concentration is very, is very, is another element… I’m going straight in on the notes not worrying about the breathing, I’m trying to get a notes right.”

P3 page 18 line 387

Participants get feedback from the tune regarding the quality of their breathing:

“if it’s clear, if it’s what I want it to be, and how I feel it should sound, and all how I know it sounds when you know a tune, you know exactly how it should sound. When I achieve that, then I know that my breathing is good”

P5 page 10 line 227

With more experience playing the harmonica, individuals commented that breathing whilst playing the harmonica felt significantly different to breathing during exertion or singing. Participants viewed this difference as beneficial, either because it was requiring more effort than singing, particularly those newer to it, or that the different breathing was providing a complimentary benefit to breathing control from singing.

“I’m about 70% of normal, but you don’t need an awful lot of power to play the harmonica from what I’ve picked up. I may be wrong, but you don’t blow in really hard. Not like blowing the trumpet where you know gotta have a lot of breath, so that’s more so, than singing, where you’ve got to perhaps use a lot more of your body. We should be using all of your body as I’ve discovered by the time (name) has got, you don’t bend your knees, you know stand up straight, don’t bend your knees, hold your stomach in, then blow it out, you know another time I’ve learned all these instructions, you make your arms flopping all the rest of it. Whereas, harmonica is, go, go for it.”

P4 page 12 line 279

“When you’re young you know you don’t think about your breathing it just happens doesn’t it, or when you can’t do it, it’s quite frightening, you know it really is, you think you know what’s happening here? Now, I now know how to control it a lot better, and how to get over these shortness of breath times, you know. And I think this has helped, the harmonica, because it pushes me, the singing as I say always sung, so it’s not too much of an effort.”

P6 page 16 line 378

Not everyone forgot about their breathing or found it easier when playing the harmonica. The participant below hadn’t been playing the harmonica for years like some of the others and found it difficult, reflecting the theme above of finding it hard to play in the beginning:

“I found the singing better really because although it might make me cough, I do that more, lungs are more open after the singing, whereas with the harmonica I feel like my chest is quite tight, so I feel the singing is doing my lungs more good really, and I feel I have a better result after singing lesson than the harmonica”

P7 page 5 line 109

### Gives you a high

Playing the harmonica is fun, it’s enjoyable and it gives people a buzz. Players are left on a high, full of positivity. The high feeling is a sense of being proud in achievement, gaining a new skill, having self-belief and being able to perform a tune in a world outside of disease, without anxiety.

“What a buzz, there’s nothing like it at all you know we come away, we usually go to the pub afterwards because it’s nice, you come away because you’re still on a high.”

P5 page 15 line 374

“When I play that, you know, you feel so comfortable in yourself you actually feel comfortable in yourself because you sitting, that, it’s like you’re in a world of your own, and that’s basically what it is. We start to feel comfortable in your own body and relax and you are more relaxed in your own body you know. And you get more confident as well. The more that you play the more confident you get”

P1 page 14 line 335

Part of being on a high is the escapism of being enthralled by the charismatic teacher.

“(Name) makes it very entertaining and at the end of the day I can get a bit of a tune out of the harmonica, all be it not very good, but it’s just it’s just good fun, and I feel that because I’m struggling too with the in breaths I feel that it’s maybe doing some good because I’m breathing out to breathe in through this, and it is, you have to make a bit more of an effort. I enjoy it, and that’s the reason why you do something.”

P6 page 6 line 121

“He’s amazing he he’s like the mad Hatter…coz he’s just in the world of his own but it’s his teaching and the way that he is with, he just gives us that good feeling the minute he comes on so we just love it”

P1 page 7 line 151

### Needing the zoom class

COVID-19 has meant that many of our social lives have been reliant on online communication with others. This was no different for individuals in this study, and discussed the necessity, advantages and disadvantages of playing the harmonica online. It became a priority in their weekly schedule.

“It’s made a huge difference to the way I live, and to my to life in general”

P5 page 2 line 41

“You’re in a group in a room it’s more, it can be more embarrassing I suppose, if you go wrong, whereas when you’re online he always says don’t worry it’s just me”

P4 page 13 line 311

“Harmonica is brilliant, and it’s, let’s keep the fact it’s keeping us alive you know, with the hope that you know we’re going to be together, we’re going to be doing it again and will be physically together again.”

P3 page 8 line 178

However other felt isolated by it and missed the genuine face to face group contact:

“I miss that banter, he was always bantering with somebody, you know, and little jokes. And what have you miss that personal contact, you know which is good. You chatter and then get told off for chattering, you know, or you’re on, and then he’ll say are you tired(name)? You know, and it’s just missed that that impromptu chatter and just being with people I think makes an awful difference. It makes really good difference. But the zoom is extremely good considering our circumstances you know. If we didn’t have all this technology we would be in terrible states, I mean I would be here I wouldn’t have spoken to anybody for nearly a year.”

P6 page 9 line 209

## Discussion

### Attuned to breathing

This qualitative analysis explored the experiences of individuals playing the harmonica with COPD. No previous investigation of harmonica playing has been explored in individuals with COPD. The main theme from the analysis was that playing the harmonica both requires, and provides a sense of breathing control. This is because it is necessary to breathe differently through the harmonica with concentration on connecting the breathing with producing sound. The altered breathing was helping to clear the chest and helping to prevent the disease worsening in lieu of access to other interventions and face to face check-ups. The sense of achievement from being able to perform a tune was empowering and left participants feeling high. Playing the harmonica appeared to be more difficult for those who had learnt over Zoom and perhaps were finding playing, and therefore breathing during the class difficult. This difference in experience between those who had played for years compared to the newcomers, in combination with reflections from experienced players offers some temporal analysis. With practice using the correct technique, players move from finding it difficult to breathe to producing a tune, being more focused on the breath with lots of concentration to using the harmonica to escape from thinking about breathing with a sense of feeling comfortable in oneself. From a phenomenological perspective temporality is part of the lived experience in any phenomenon, and with harmonica playing online, it offered the participants hope, in looking forward to being-with-others again.

### A social need

Participants reported the importance of the Zoom class in their week, enabling them to be with others. It also enabled more of a perceived personalised interaction with the group leader, with reduced potential for social embarrassment. However, the buzz, the vibe, or the banter was lost, which was part of the attraction when joining the group face-to-face.

SLH group participation has also been shown to successfully adapt to online delivery with positive health outcomes (19), and the social lifeline the Zoom class is offering participants playing the harmonica, may continue for some time. With COVID-19 still present, many individuals with respiratory disease may be wary of returning to groups. Indeed, the majority of Individuals with respiratory disease are keen to continuing social distancing and mask wearing(20), both of which make playing harmonica in a group difficult. There has been a reported 50% reduction in COPD admissions during COVID-19(21). Providing Zoom-based options for group-based interventions may not only provide the social lifeline described above, but also reduce infection risk.

### An adjunct

The resistance of the harmonica at the mouth means that increased pressures are likely needed to be generated on the inhalation and exhalation to trigger and maintain flow for the breath. From a physiotherapy perspective the harmonica appears to act as an inspiratory muscle trainer and as a positive expiratory pressure device, performing a technique akin to pursed-lip breathing. Participants repeatedly commented that drawing was harder than blowing into the harmonica and that with blowing there was a sense of release or ease, particularly by those with more experience. Further mechanistic studies are warranted to investigate the potential of the harmonica as an airway clearance adjunct

## Strengths and Limitations

This study provides in-depth experiences of individuals who are both very experienced and relatively novice at playing the harmonica. This enabled rich and varied accounts of the lived experiences of harmonica playing. The open-ended nature of questioning in combination with the fact that AL was previously unknown to the participants enabled individuals to be forthright in verbalising not only perceived benefits but detailing difficulties experienced. However, the sample of participants was unique. Many had combined previous experiences of both participating in PR and SLH groups. This could limit the transferability of the findings to those individuals who haven’t previously participated in group-based interventions. However, these combined experiences enabled a greater reflection of the meaning of harmonica playing in context with other interventions which is valuable for clinicians and patients to understand. Further feasibility studies and then adequately powered randomised controlled trials investigating social and clinical outcomes of face-to-face, and online harmonica groups for individuals with CRD is now warranted.

## Conclusion

Playing the harmonica is a novel intervention for individuals living with CRD. This study investigated the lived experiences of individuals playing the harmonica in a group of others living with CRD. Participants experienced breathing differently because of playing the harmonica. Breathing with the harmonica, particularly during inspiration was hard at first, but with experience, playing the harmonica offered control of their breathing, in a fun, social activity which made them feel they were preventing their disease from worsening. Further mechanistic and randomised controlled trials are warranted to determine the clinical value of harmonica playing when living with COPD.

## Data Availability

Data are available upon reasonable request.

## Acknowledgements

Thank you to all the participants who agreed to participate and generously volunteer their time on this study.

## Author Contributions

The authors meet criteria for authorship as recommended by the International Committee of Medical Journal Editors. AL, JC, CS and JW designed the study. AL recruited participants, performed the interviews, transcription and analysed the data. JM (student physiotherapist) supported transcription within his education. JW and CS supported recruitment and CS led the harmonica groups. AL wrote the initial draft of the manuscript which all authors reviewed and contributed towards the final draft.

## Data Availability Statement

Data are available upon reasonable request.

