## Supplementary material for "Playing the Harmonica with Chronic Obstructive Pulmonary Disease. A qualitative study": online supplement

### Semi-structured interview guide

Hello, my name is….Thank you for your time today and agreeing to participate in our study. I will summarise the study according to information already provided in the Participant Information Sheet and inform you once more of the potential risks of participating. After that I can answer any further questions you may have and we can go through the consent process.

**Description of study, potential risks and consent procedure**

**Questions**

1] So, how are you today?/What sort of day are you having today?

2] Tell me a little about yourself (prompts including how their see themselves, roles, identity etc)

3) If you think about your COPD, paint a picture for me verbally of what is going on in your lungs

- and with your breathing

4] Tell me more about your life living with COPD before using the harmonica

- What did a typical day/week look like?
- Living with a long-term condition?
- Living with others?
- Your life at home?
- Your involvement in activities and/ or groups outside of home?
- Plans for your future?

4] Can you give me an example of when you have used the harmonica recently?

- What was the experience like?
- What sort of things are you thinking about?
- How did it make your feel? (psychological, emotional, physical and social impact)
- Describe how it made your breathing feel?
- How does your breathing and harmonica playing work together?
- How does the musicality of playing harmonica effect you?

5) What made you join the online group?

- What happens during a harmonica session?
- What does having a group of you playing together mean to you?

6) Has being a member of the online group influenced your priorities or the way you engage with life outside the online group?
7) Describe an experience of how you have used the harmonica to manage your COPD specifically

- Symptoms
- Mood
- Physical activity
- How does playing the harmonica compare with other activities you may do to manage your breath?

9] Now let’s talk about your thoughts and plans about the future…

- For you in general
- Playing the harmonica
- Managing your COPD

10] What else do you think is important to consider at the moment regarding harmonica playing, being a member of an online group, or living with your condition?

11) Is there anything else you would think valuable to talk about or ask others living in a similar situation?

Thank you for your time. The research team will be in contact with the leaders of your harmonica group when the research findings are ready to be shared with you.

Have a good rest of your day.

### Further example quotes

| Theme | Quotes |
| --- | --- |
| Hard in the beginning | “it's like nothing is happening it's like having a hole that's filled up I said when you breathe there's not enough room to be able to to pull that in. You know and and it's learning how to be able to release that a little bit, I think it's just a use of muscles that we perhaps forgot forgotten how to use or have never used properly because probably over the years it was a struggle so it's your muscles give up don't know if you don't get used and I think that's what it's having to do is to release that tension in the muscle.”  P5 page 4 line 88  “I did find it hard at first because with the breathing with the harmonica it's from the throat so at first when you learning you're thinking it's coming from your diaphragm so you doing like you do at singing and then obviously with (name) is teaching but he does it in a way we do it in separate stages so we get to learn how to breathe properly, when to breathe, and then I found I found it was easier.”  P1 page 1 line 37  “drawing is actually much harder than the blowing  and to keep that going for some length of time it is really quite difficult”  P7 page 4 line 79  “when you're blowing out you you've already taken a breath in before you start playing and when you blow out it's it's quite simple when you drawback it's catching you in your throat, and then probably getting mixed up in in the phlegm in your throat or whatever, I dont know. But it's quite hard especially if you have to draw a note and hold it for longish time you know that's the hard part and especially if you have to draw throughout 4 notes in a row that's even worse”  P2 page 4 line 72 |
| Holding the condition | “carry on sorting my chest out or keeping it stable as much as it can be”  P4 page 15 line 353  “you're there to work your lungs etcetera with the various different exercises so once you get used to that then you you're also starting to use those exercises you know you know the breathing in, breathing out, holding it, all sorts of things like that, you know, deal with the mucus, deals with the coughing.”  P3 page 4 line 89  “I found that it is, hold it, hold it, and so I can deal with it, and work with it, and do all the things I've done with it, you know because at the end of the day with something I'm very proud of”  P3 page 10 line 227  “Quite a few of us have coughing as soon as we start you know because it does help to clear the mucous in your lungs and it does help”  P5 page 3 line 68 |
| Breathing control | “I was blowing and drawing from my chest and diaphragm and that was quite hard and then one of the other people and (name) both said don't blow from there, blow from your throat take it nice and easy, nice and gentle, and I've been doing that ever since and I find it quite easy and it is helping, I do feel my COPD is fairly well under control…”  P2 page 2 line 37  “I used to get breathless even just talking, I used to get breathless, now playing harmonica and I don't feel breathless at all when I’m finished.”  P2 page 2 line 48  “It's made me very conscious of my breathing, how I'm breathing, and what I can do to help in the breathing situation”  P6 page 16 line 360  “So I’ll start with quite a strong sort of note, but it'll disappear on me because I haven’t got enough air to keep it up”  P7 page 9 line 206  “It just sends you into a different world you know, and you're not even thinking of your problems or anything and you’re just sitting there, and you know you’re thinking oh I can play this song now and you're not short of breath, yeah I mean it, you just feel good about it.”  P1 page 10 line 220 |
| Gives you a high | “Thinking about getting the right notes, quite a hard question really, it's a calming influence and you're concentrating on what you doing and it gives you a high you know because you can pick it up anytime and just wonder about.”  P2 page 7 line 156  “Confidence comes up, well-being comes up satisfaction, belief, togetherness all these things come around it let alone the breathing part.”  P3 page 12 line 272  “His way of dealing with all of it is really uplifting he's you're very positive he's funny, you can't come away from a lesson with him feeling miserable it's as simple as that.”  P5 page 15 line 354  “You come away with a smile on your face think it's been the instructor yes is absolutely brilliant is absolutely bonkers but he's absolutely brilliant even when it was face-to-face, always come away smile on my face, yeah really really lifts you”  P8 page 7 line 160 |
| Needing the zoom class | “The zooming comes first it does come yes yeah well we look forward to it.”  P1 page 12 line 278  “I have been locked down or shielding or whatever since last March you know you know my cars done 1000 miles in a year you know it's it's ridiculous isn't it. Yes to concentrate on keeping yourselves active in the house doing, rather than just sitting about moaning, and groaning. Just do little things at a time. And this singing and harmonica playing is ideal thing for us at the moment, we, we, we wouldn't really be existing without the zoom class you know because for what else could we do I mean and I'm not well you know about my age I I'm not very technically minded but I’ve had to learn and its Zoom has been great a great asset to us now can't be without it because in in in the classes we have made a lot of friends and we are always great seeing them together”  P2 page 6 line 129  “I'm not sure how confident I would feel about being part of the live group.  I will probably try it and see how I feel because to be honest when you're in a live class you can see that other people have the same problems  sometimes makes you feel better in which case I would continue and if it was too much of a struggle then obviously I wouldn’t continue it.”  P7 page 6 line 136 |
